# Home-based transcranial direct current stimulation for major depressive disorder: 6-month follow-up from a randomised sham-controlled trial

**DOI:** 10.1101/2024.11.21.24317690

**Authors:** Rachel D. Woodham, Sudhakar Selvaraj, Nahed Lajmi, Harriet Hobday, Gabrielle Sheehan, Ali-Reza Ghazi-Noori, Peter J. Lagerberg, Rodrigo Machado-Vieira, Jair C. Soares, Allan H. Young, Cynthia H.Y. Fu

**Affiliations:** Department of Psychology, University of East London, London, UK; Center of Excellence on Mood Disorders, Faillace Department of Psychiatry and Behavioral Sciences, McGovern Medical School, University of Texas Health Science Center at Houston, Houston, TX, USA; Intra-Cellular Therapies Inc., USA; Centre for Affective Disorders, Institute of Psychiatry, Psychology and Neuroscience, King’s College London, London, UK; National Institute for Health Research Biomedical Research Centre at South London and Maudsley NHS Foundation Trust, King’s College London, London, UK; South London and Maudsley NHS Foundation Trust, Bethlem Royal Hospital, Beckenham, UK

**Keywords:** transcranial direct current stimulation, neuromodulation, non-invasive, major depression, long term outcome

## Abstract

Transcranial direct current stimulation (tDCS) is a potential home-based treatment for major depressive disorder (MDD). In our randomised controlled trial, a fully remote, 10-week course of home-based active tDCS showed greater clinical efficacy as compared to sham tDCS in a double-blind randomised controlled trial (RCT). tDCS was administered at 2mA for 30-minute sessions in a bifrontal montage with anode over the left dorsolateral prefrontal cortex (DLPFC) and cathode over the right DLPFC. Following the 10-week blinded trial, all participants could receive active tDCS in a 10-week open label phase. Participants who had completed the open-label treatment were invited for additional long term follow-up visits over 6 months. Participants were allowed to keep their tDCS device and could choose to continue stimulation during the follow-up period. In the follow-up phase, 42 participants (27 women) attended at least one visit. Ongoing use of the tDCS device was 59% at 3 months and 55% at 6 months during the follow-up period. The clinical response rate was 64% at 3 months and 76% at 6 months. Participants who had shown a clinical response after open label treatment maintained a response rate of 90% at 6 months. In summary, high rates of clinical response were maintained in 6-month follow-up whether or not participants continued to use the tDCS device.

## 1. Introduction

Major depressive disorder (MDD) is a leading cause of disability worldwide (World Health Organization, 2017) and is characterized by a prolonged low mood or an inability to experience feelings of pleasure that is associated with impairments in cognition, psychomotor functioning and disturbances with sleep, appetite and energy levels. Current first line treatments are antidepressant medications and psychological therapies. However, over a third of people with MDD do not achieve a full clinical remission despite full treatment trials (Cuijpers et al., 2014; Rush et al., 2006).

tDCS is a non-invasive form of brain stimulation which modulates cortical tissue excitability by applying a weak (0.5-2 mA) direct current via electrodes (Woodham et al., 2021). tDCS does not directly generate action potentials in neuronal cells, in contrast to repetitive transcranial magnetic stimulation (rTMS) (Mutz et al., 2019). Rather, tDCS modifies the membrane polarity of neurons and, thus, their threshold for action potential generation (Nitsche and Paulus, 2000). For the treatment of MDD the anode electrode is typically placed over the left dorsolateral prefrontal cortex (DLPFC) and cathode over the right DLPFC, suborbital or frontotemporal region (Mutz et al., 2019). Meta-analyses have reported clinical benefits of active as compared to sham tDCS in MDD (Mutz et al., 2019).

Long term follow-up assessments at 6 months have observed relapse rates from 26 - 53% in MDD and bipolar depression following a course of tDCS, ranging from 3 - 6 weeks and ongoing treatment sessions over 6 months (Martin et al., 2013; Valiengo et al., 2013; Apraicio et al., 2019). Razza et al. (2021) meta-analysis reported a moderate to large improvement of tDCS treatments effects in the 6-month follow-up period as compared to the end of trial measure in interventional studies (Razza et al., 2021). In our clinical trial of 174 participants with major depressive disorder (MDD), a significantly greater improvement in clinician-rated depressive symptoms, self-reported depressive symptoms, clinical response and remission rates were observed in the active as compared to sham group at 10 weeks (Woodham et al., 2024). Following unblinding, all participants were given the option to receive active tDCS for a further 10 weeks (Woodham et al., 2024). However, whether the effects are maintained in a longer term is unclear.

The present study investigates the long-term efficacy, safety and acceptability of home-based self-administered tDCS treatment in a 6-month observational follow-up of participants who had received a 10-week course of home-based tDCS in a phase 2 randomized controlled trial (Woodham et al., 2024).

## 2. Material and Methods

### 2.1. Trial design

Ethical approval was provided by the South Central-Hampshire B Research Ethics Committee, UK (ref. 22/SC/0023) and WIRB-Copernicus Group International Review Board, USA (ref. 1324775). All participants provided written informed consent to participate in the clinical trial and follow-up study. The double-blind, placebo-controlled, randomized, superiority trial of home-based tDCS in MDD (ClinicalTrials.gov NCT05202119) was conducted in England and Wales, UK, and Texas, USA, at University of East London and University of Texas Health Science Center at Houston, respectively. The long term follow-up was conducted at the UK study site as the trial duration required extended observation, ethics approval had been obtained in the UK site and participants were able to keep the tDCS device.

All study visits were completed by videoconference using Microsoft Teams. The trial consisted of a 10-week blinded treatment phase, where participants were randomly assigned to receive either active or sham tDCS, followed by 10-week open label phase. Active stimulation was 2mA direct current stimulation for 30 minutes with gradual ramp up over 120 seconds at the start and ramp down over 15 seconds at the end of each session. Sham stimulation had an initial ramp up from 0 to 1 mA over 30 seconds then ramp down to 0 mA over 15 seconds and repeated at the end of session. Anode and cathode were over the left and right dorsolateral prefrontal cortices, respectively. The blinded phase of the trial consisted of 5 tDCS sessions per week for 3 weeks followed by 3 sessions per week for 7 weeks. The open label phase consisted of active tDCS sessions for all participants. Participants in the initial active tDCS treatment arm could complete 3 sessions per week for 10 weeks, and participants who had been in initial sham tDCS treatment were offered the active tDCS stimulation schedule, 5 sessions per week for 3 weeks then 3 sessions per week for 7 weeks. A full description of trial design and results has been published elsewhere (Woodham et al., 2024).

Participants who had completed blinded and open label phases of the trial were invited to participate in follow-up visits at 3-months and 6-months after the 20-week phase of the trial. During the follow-up period, participants were not under any instruction regarding device use. The final follow-up was conducted on January 26, 2024.

### 2.2. Participants

174 participants were enrolled (mean age 37.63 ± 11.00 years, 120 women), with MDD in current depressive episode based on Diagnostic and Statistical Manual of Mental Disorders, Fifth Edition (DSM-5) criteria (American Psychiatric Association, 2013) by structured clinical assessment, Mini-International Neuropsychiatric Interview (MINI; Version 7.0.2) (Sheehan et al., 1998), having at least a moderate severity of depressive symptoms, as measured by score 16 or more on 17-item Hamilton Rating Scale for Depression (HDRS) (Hamilton, 1960). All participants were under the care of a GP and could be treatment-free, or taking stable antidepressant medication or in psychotherapy for at least 6 weeks prior to enrolment and agreeable to maintaining same treatment throughout the trial. Exclusion criteria included: having treatment resistant depression, defined as inadequate clinical response to two or more trials of antidepressant medication at an adequate dose and duration; significant suicide risk based on Columbia Suicide Severity Rating Scale (C-SSRS) Triage and Risk Identification Screener (Posner et al., 2011); comorbid psychiatric disorder; taking medications that affect cortical excitability (e.g., benzodiazepines, epileptics); and contraindications to tDCS. Following the 10-week RCT, 141 participants continued in the 10- week open label phase (100 UK, 41 USA).

### 2.3. Assessments and outcomes

Follow-up assessments were performed at 3-months and 6-months following the 20-week end of the open-label treatment phase. Depressive severity was measured by clinician-rated scales, HDRS, Montgomery-Åsberg Depression Rating Scale (MADRS) (Montgomery and Åsberg, 1979), suicide ideation with Columbia-Suicide Severity Rating Scale (C-SSRS) (Posner et al., 2011); mania symptoms with Young Mania Rating Scale (YMRS) (Young et al., 1978); anxiety symptoms with Hamilton Anxiety Rating Scale (HAMA) (Hamilton, 1959), and quality of life with EQ-5D-3L (Brooks, 1996; “EuroQol - a new facility for the measurement of health-related quality of life,” 1990; Rabin and de Charro, 2001), which has five dimensions: mobility, self-care, usual activities, pain and discomfort, and anxiety and depression, and three severity levels. Adverse events were assessed using the tDCS Adverse Events Questionnaire (AEQ) (Brunoni et al., 2011). Treatment acceptability was assessed by our treatment acceptability questionnaire (TAQ) (Rimmer et al., 2024; Woodham et al., 2022). Clinical response was assessed by a HDRS score reduction of at least 50% relative to the baseline HDRS score, and clinical remission was a HDRS score of 7 or less.

### 2.4. Statistical Analysis

An intention to treat analysis was completed, using a last observation carried forward (LOCF) method for missing data on clinical assessments. Four mixed ANOVAs were conducted, with original treatment arm as the between-subjects variable, HDRS, MADRS, HAMA and EQ-5D-3L total scores were the dependant variables and assessment time-point was the within-subjects factor, with four levels including end of blinded treatment period (*t*1), end of open label treatment period (*t*2), 3-month follow-up (*t*3) and 6-month follow-up (*t*4).

Two mixed ANOVAS were completed using data from participants who had attended each of the two follow-up visits to explore if there was any interaction between continued tDCS use during the follow-up period and depressive symptoms. Both ANOVAs included tDCS use during that follow-up period as the between-subjects variable, HDRS total score was the dependant variable, and assessment time point was the within-subjects factor with two levels. For the first ANOVA these were the end of open label treatment period (*t*1) and the 3- month follow-up (*t*2), and for the second ANOVA they were and the 3-month follow-up (*t*1) and the 6-month follow-up (*t*2).

Statistical analyses were conducted using IBM SPSS Statistics (version 29.0.1.0). All analyses were two tailed and significance value of *p* = 0.05 was set. Greenhouse-Geisser correction was applied if Mauchley’s assumption of sphericity was violated. Post hoc pairwise comparisons with Bonferroni corrections were conducted.

## 3. Results

### 3.1. Clinical assessments

A total of 174 MDD participants were enrolled and randomised to active (n=87) or sham tDCS (n=87) treatment arms. From the 10-week RCT phase, 149 MDD participants completed the week 10 end of treatment assessment. From the next 10-week open-label treatment phase, 111 MDD participants completed the week 20 assessment (77 UK, 34 USA) (Figure 1).

**Figure 1.**
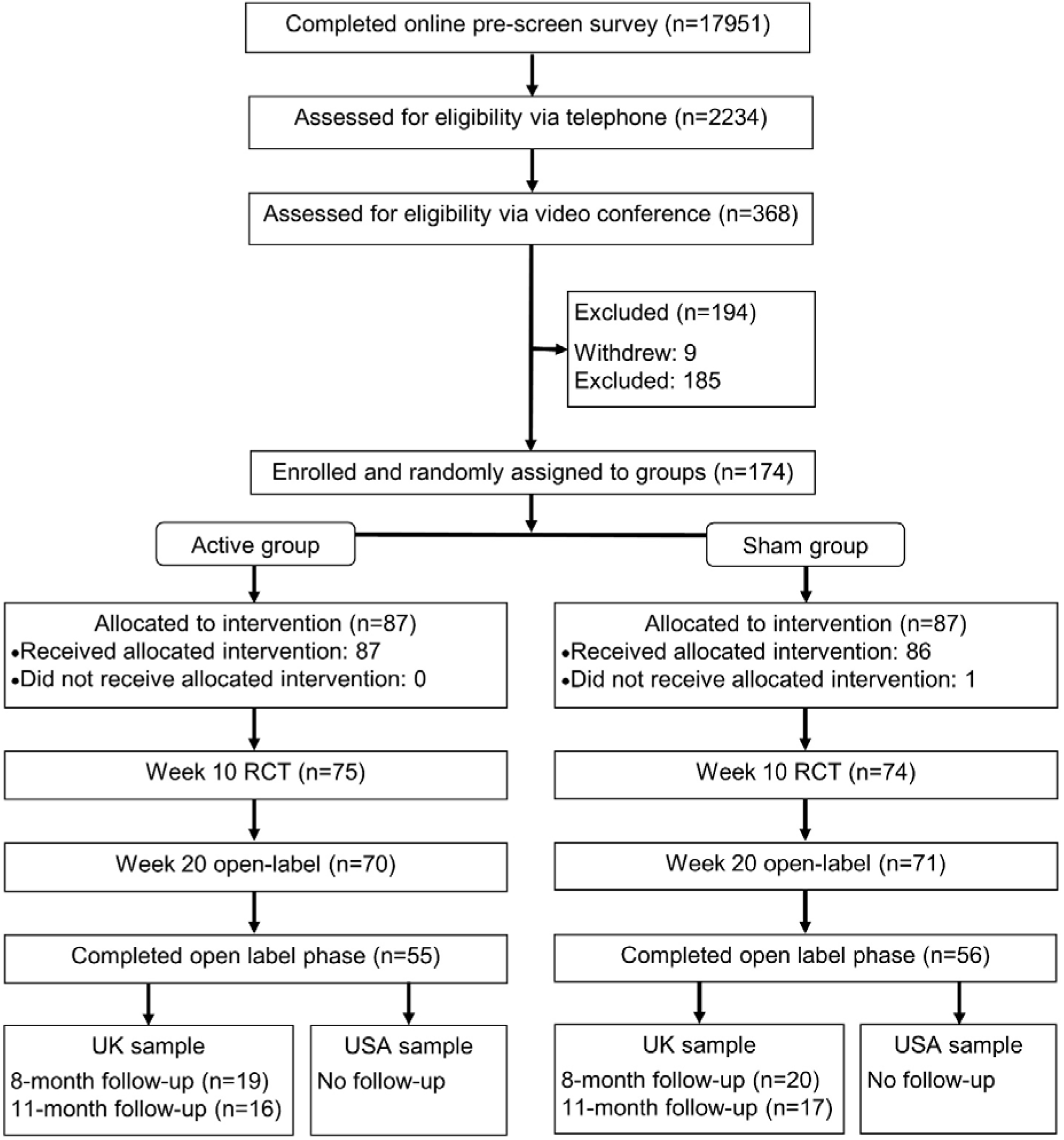
Flow chart of the blinded phase, open label phase and follow-up visits of the tDCS for major depression at home study (Empower study).

In the UK sample, 77 participants had completed the combined 20-week RCT and open-label phase.

From participants who had been in the active treatment arm (n = 35), mean HRSD was 8.51 ± 5.66 at week 20. Clinical outcomes were: treatment response (n = 21; 60.0%), who completed an average 17.68 ± 9.66 active tDCS sessions (range 2-30 sessions) in 10-week open label phase and total 50.92 ± 12.32 active tDCS sessions (range 23-66 sessions) over 20 weeks; no treatment response (n = 14; 40.0%), consisting of participants (n = 11) who completed average 14.02 ± 7.61 active tDCS sessions in the open label phase, and participants (n = 3) who did not continue with any sessions.

From participants who had been in the sham treatment arm (n = 42), mean HRSD was 9.54 ± 5.22 at week 20. Clinical outcomes were: treatment response (n = 25; 59.5%), who completed an average 28.00 ± 9.30 active tDCS sessions (range 1 - 38 sessions) in 10- week open label phase and an average 33.94 ± 3.17 sham sessions (range 25 - 37 sessions) over the 10 week blinded phase; no treatment response (n = 17; 40.5%), consisting of participants (n = 17) who completed average 24.30 ± 12.69 active tDCS sessions in open label phase, and participants (n = 0) who did not continue with any sessions.

Additional follow-up visits were conducted after 3- and 6-months (at 8- and 11-months following randomisation). 42 MDD participants (27 women) (54.5% from week 20 in UK site) attended at least one follow-up visits (mean age 38.07 ± 10.93 years; mean baseline HDRS 18.21 ± 2.28) (Table 1). Concurrent treatments at baseline were: antidepressant medication (n=27), combination of antidepressant medication and psychotherapy (n=4) (antidepressant medication duration: range 6 weeks - 30 years, psychotherapy duration: range 13 - 100 weeks), and being treatment-free (n=15). Treatment arm allocation had been active tDCS (19 participants; 11 showed a clinical response at week 10) and sham tDCS (23 participants; 10 showed clinical response at week 10).

**Table 1.** Baseline demographic and clinical characteristics of participants at the start of the trial and week 20 outcomes.

| Characteristic | Total<br>(N=42) | Active<br>(N=19) | Sham<br>(N=23) |
| --- | --- | --- | --- |
| Age | 38.07 ± 10.93 | 36.68 ± 11.96 | 39.22 ± 10.12 |
| Gender |  |  |  |
| Female | 27 (64) | 11 (58) | 16 (70) |
| Race |  |  |  |
| Asian | 3 (7) | 2 (11) | 1 (4) |
| Black or African American | 1 (2) | 0 (0) | 1 (4) |
| White | 37 (88) | 16 (84) | 21 (91) |
| Other | 1 (2) | 1 (5) | 0 (0) |
| Educational □ Level |  |  |  |
| Secondary School | 2 (5) | 2 (11) | 0 (0) |
| College | 11 (26) | 2 (11) | 9 (39) |
| Bachelor's or Professional Degree | 16 (38) | 8 (42) | 8 (35) |
| Master's or Doctoral Degree | 13 (31) | 7 (37) | 6 (26) |
| Age of onset of MDD | 22.93 ± 9.70 | 19.74 ± 7.34 | 25.57 ± 10.70 |
| Previous number of episodes | 2.5 (0, 5) | 1 (0, 4.5) | 4 (0.5, 5) |
| Previous number of suicide attempts | 0 (0, 0) | 0 (0, 0) | 0 (0, 0) |
| First episode MDD | 14 (33) | 8 (42) | 6 (26) |
| Clinical ratings |  |  |  |
| HDRS baseline | 18.21 ± 2.28 | 18.16 ± 1.92 | 18.26 ± 2.58 |
| HDRS week 20 response | 28 (67) | 13 (68) | 15 (65) |
| HDRS week 20 remission | 23 (55) | 12 (63) | 11 (49) |
| MADRS baseline | 24.17 ± 4.25 | 24.05 ± 3.36 | 24.26 ± 4.94 |
| HAMA baseline | 14.81 ± 4.59 | 14.74 ± 4.04 | 14.87 ± 5.08 |
| YMRS baseline | 2.43 ± 1.74 | 2.37 ± 2.01 | 2.48 ± 1.53 |
| EQ-5D-3L baseline | 0.73 ± 0.18 | 0.71 ± 0.18 | 0.75 ± 0.18 |
| Total number of active tDCS sessions completed | 39 ± 14.56 | 49 ± 14.46 | 30 ± 8.02 |
| Range of active tDCS sessions completed | 9 - 66 | 23 - 66 | 9 - 39 |
| Antidepressant medication during trial | 27 (64) | 14 (74) | 13 (57) |
| Selective serotonin reuptake inhibitor | 22 (52) | 13 (68) | 9 (39) |
| Non-selective monoamine reuptake inhibitor | 1 (2) | 0 (0) | 1 (4) |
| Other antidepressant medications | 5 (12) | 2 (11) | 3 (13) |
| Combination psychotherapy and antidepressant medication | 4 (10) | 1 (5) | 3 (13) |
| No antidepressant medication or psychotherapy during trial | 15 (36) | 5 (26) | 10 (44) |

Characteristics are presented for participants who attended at least one of the two follow-up visits at month 8 or month 11. Categorical variables are presented as number of participants with percentage in parentheses for gender, race, educational level, first episode MDD, antidepressant medication and individual medications, individual psychotherapy during trial and No antidepressant or psychotherapy during trial. Mean values are presented with '±' standard deviation values. Previous number of episodes and previous number of suicide attempts are presented as median with interquartile range in parenthesis. HDRS, Hamilton Rating Scale for Depression; MADRS, Montgomery-Åsberg Depression Rating Scale; HAMA, Hamilton Anxiety Rating Scale; YMRS, Young Mania Rating Scale; EQ-5D-3L, quality of life measure (https://euroqol.org). One participant in the active group was taking a combination of one other antidepressant medication and one selective serotonin reuptake inhibitor.

At month 8, participants completing follow-up (n = 39), consisting of participants maintaining tDCS sessions (n = 23), number of sessions: 9 – 33 in 3 months, and participants who discontinued sessions (n = 16). Concurrent treatments were: antidepressant medication (n = 25), standalone psychotherapy (n = 1), combination medication and psychotherapy (n = 1), and being treatment-free (n = 13) (Table 2). Based on HDRS ratings, mean was 7.50 ± 4.80, and clinical outcomes were: treatment response (n = 25; 64%), remission (n = 22; 56%). Based on MADRS ratings, mean was 10.11 ± 6.86, and clinical outcomes were: treatment response (n = 26; 67%), remission (n = 24; 62%), HAMA mean 7.53 ± 5.19, and EQ-5D-3L mean 0.84 ± 0.17.

**Table 2.** Depression treatments at month 8 and month 11 follow-up assessments.

| **Month 8** | **Total**<br>**(N=39)** | **Week 20 responders**<br>**(n=26)** |
| --- | --- | --- |
| Antidepressant medication | 25 (64) | 14 (54) |
| Selective serotonin reuptake inhibitor | 18 (46) | 10 (38) |
| Non-selective monoamine reuptake inhibitor | 1 (3) | 0 (0) |
| Other antidepressant medications | 6 (15) | 4 (15) |
| Standalone individual psychotherapy | 1 (3) | 1 (4) |
| Combination psychotherapy and antidepressant medication | 1 (3) | 1 (4) |
| Treatment free | 13 (33) | 11 (42) |
| Continued regular tDCS | 23 (59) | 18 (69) |
| Once per week | 3 (8) | 3 (12) |
| Twice per week | 14 (36) | 10 (38) |
| Once or twice per week | 2 (5) | 2 (8) |
| Reset stimulation to 5 times per week | 4 (10) | 3 (12) |
| **Month 11** | **Total**<br>**(N=33)** | **Week 20 responders**<br>**(n=21)** |
| Antidepressant medication | 20 (61) | 10 (48) |
| Selective serotonin reuptake inhibitor | 14 (42) | 6 (29) |
| Non-selective monoamine reuptake inhibitor | 1 (3) | 0 (0) |
| Other antidepressant medications | 5 (16) | 4 (19) |
| Standalone individual psychotherapy | 3 (8) | 3 (12) |
| Combination psychotherapy and antidepressant medication | 1 (3) | 1 (5) |
| Treatment free | 10 (30) | 8 (38) |
| Continued regular tDCS | 18 (55) | 14 (67) |
| Once per week | 1 (3) | 1 (5) |
| Twice per week | 13 (40) | 10 (48) |
| Once or twice per week | 2 (6) | 2 (10) |
| Reset stimulation to 5 times per week | 2 (6) | 1 (5) |

tDCS, transcranial direct current stimulation. Categorical variables are presented as number of participants with percentage in parentheses. At month 8 one participant had started taking antidepressant medication, 22 participants were taking the same antidepressant medication as before, one had changed antidepressant medication, two participants had stopped their antidepressant medications and one participant had stopped one of their medications. One participant had continued psychotherapy and one had started psychotherapy during the 3- month follow-up period. At month 11, one participant was taking the same antidepressant medication as during the clinical trial, 19 participants were taking the same antidepressant medication as they were at month 8, one participant had stopped taking antidepressant medication and one participant had a mood stabiliser added as a combination medication. No participants had started antidepressant medication that were not taking it previously. Two participants had continued psychotherapy and two had started during the follow-up period. One participant in the sham group was taking a combination of a non-selective monoamine reuptake inhibitor and a mood stabiliser at month 11. During the 3-month follow-up period to month 8, 5 participants were not stimulating for the full 12 weeks (range 3-8 weeks). During the follow-up to month 11 7 participants were not stimulating for the full 12 weeks (range 1.5 weeks – 10 weeks).

At month 11, participants completing follow-up (n = 33), consisting of 18 participants maintaining tDCS sessions (n = 18), number of sessions: 12 - 28 in 3 months, and participants who discontinued sessions (n = 15). Concurrent treatments were: antidepressant medication (n = 20), standalone psychotherapy (n = 3), combination medication and psychotherapy (n = 1), and being treatment-free (n = 10) (Table 2). Based on HDRS ratings, mean was 7.16 ± 5.15, and clinical outcomes were: treatment response (n = 25; 76%), clinical remission (n = 21; 64%). Based on MADRS ratings, mean was 8.97 ± 6.60, and clinical outcomes were: treatment response (n = 25; 76%), clinical remission (n = 24; 73%), HAMA mean 7.06 ± 5.12, and EQ-5D-3L mean 0.85 ± 0.17.

There was a significant main effect of time in HDRS score (*F*(2.14, 85.56) = 9.30, *p* < 0.001), MADRS score (*F*(2.28, 91.13) = 9.52, *p* < 0.001) and HAMA score (*F*(2.47, 98.84) = 5.08, *p* = 0.002). Pairwise comparisons showed significant improvements in HDRS scores from week 10 to week 20 (*p* = 0.002), from week 10 to month 8 (*p* < 0.001), and from week 10 to month 11 (*p* = 0.003). There was no significant change in scores over time from week 20 to month 8 (*p* = 1.00), from week 20 to month 11 (*p* = 1.00), nor from month 8 to month 11 (*p* = 1.00). A similar pattern of results was found for MADRS and HAMA scores. There were no interaction effects between time and original treatment arm in HDRS score (*F*(2.14, 85.56) = 1.29, *p* = 0.28) (Table 3, Figure 2), MADRS score (*F*(2.28, 91.13) = 1.08, *p* = 0.35) (Table 3, Figure 3) or (HAMA score *F*(2.47, 98.84) = 0.90, *p* = 0.43) (Table 3, Figure 4). Neither a main effect of time (*p* = 0.86), nor a time by group interaction effect (*p* = 0.17), was observed in EQ-5D-3L scores.

**Figure 2.**
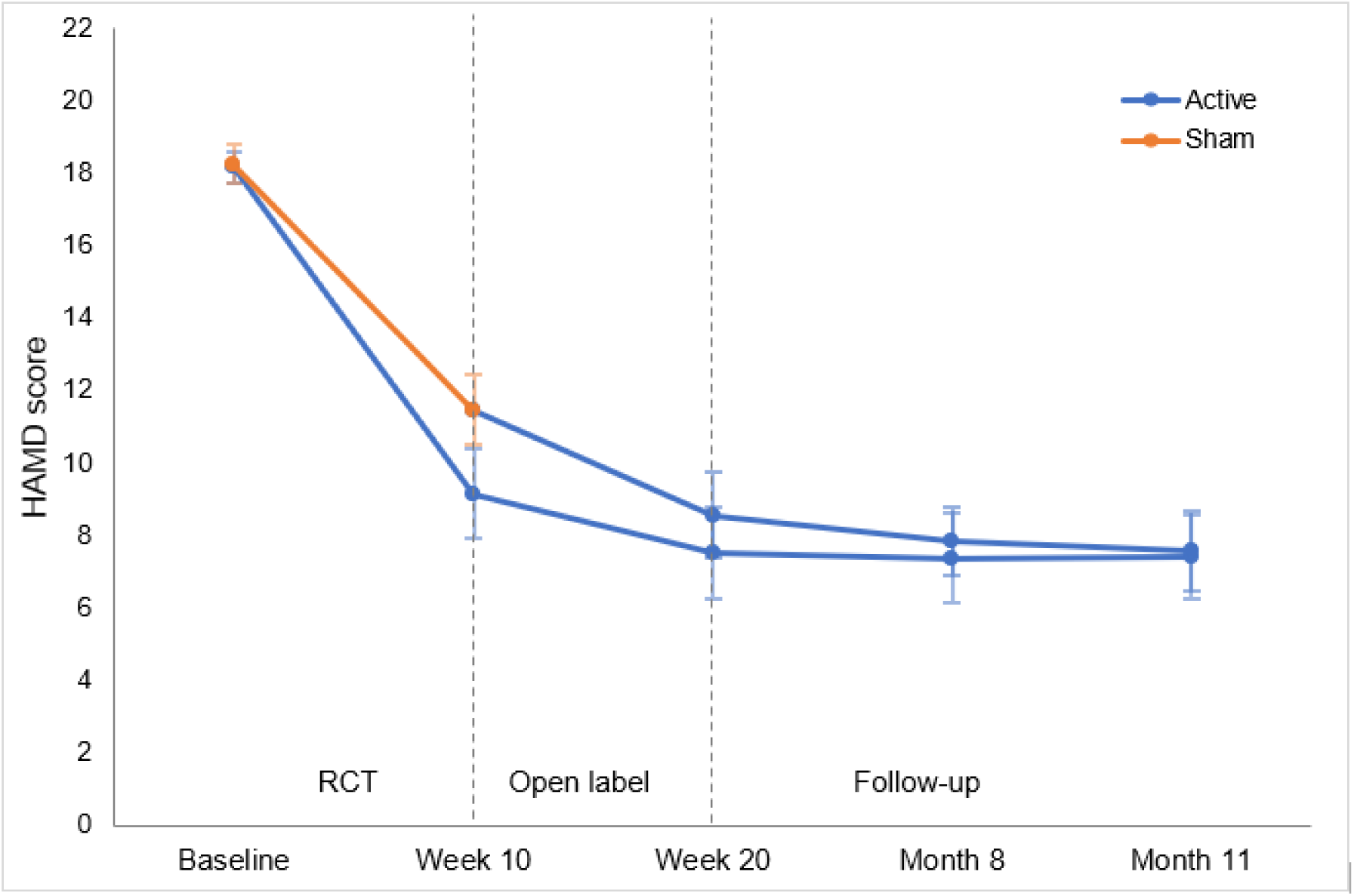
Mean Hamilton Rating Scale for Depression (HDRS) total scores for patients who were allocated to the active or sham treatment group at each assessment time point from the end of the blinded phase of the trial to the month 11 follow-up. Error bars represent standard deviation. Number of participants in the original active group (n=19) and in sham group (n=23).

**Table 3.** Clinical rating scale scores during the trial period and at follow-up, intention to treat analysis.

| Clinical ratings | Week 10 |  | Week 20 |  | Month 8 |  | Month 11 |  |
| --- | --- | --- | --- | --- | --- | --- | --- | --- |
|  | Active | Sham | Active | Sham | Active | Sham | Active | Sham |
| HDRS | 9.16 ± 5.41 | 11.48 ± 4.57 | 7.53 ± 5.43 | 8.57 ± 5.64 | 7.37 ± 5.36 | 7.87 ± 4.50 | 7.42 ± 5.00 | 7.57 ± 5.30 |
| MADRS | 12.21 ± 8.48 | 15.57 ± 7.48 | 9.68 ± 7.72 | 11.74 ± 8.26 | 9.63 ± 7.56 | 10.78 ± 6.61 | 9.42 ± 6.82 | 9.78 ± 6.80 |
| HAMA | 8.95 ± 5.80 | 10.57 ± 5.08 | 6.58 ± 5.63 | 8.74 ± 5.30 | 7.16 ± 5.46 | 7.91 ± 5.95 | 7.58 ± 4.87 | 7.70 ± 5.72 |
| YMRS | 1.37 ± 1.30 | 2.17 ± 1.83 | 1.05 ± 1.31 | 1.61 ± 1.41 | 0.95 ± 1.03 | 1.39 ± 1.70 | 0.79 ± 0.79 | 1.04 ± 1.30 |
| EQ-5D-3L | 0.83 ± 0.20 | 0.82 ± 0.16 | 0.84 ± 0.14 | 0.84 ± 0.19 | 0.80 ± 0.21 | 0.88 ± 0.11 | 0.86 ± 0.17 | 0.84 ± 0.17 |

Mean values are presented with '±' standard deviation values. HDRS, Hamilton Rating Scale for Depression; MADRS, Montgomery-Åsberg Depression Rating Scale; HAMA, Hamilton Anxiety Rating Scale; YMRS, Young Mania Rating Scale; EQ-5D-3L, quality of life measure (https://euroqol.org).

**Figure 3.**
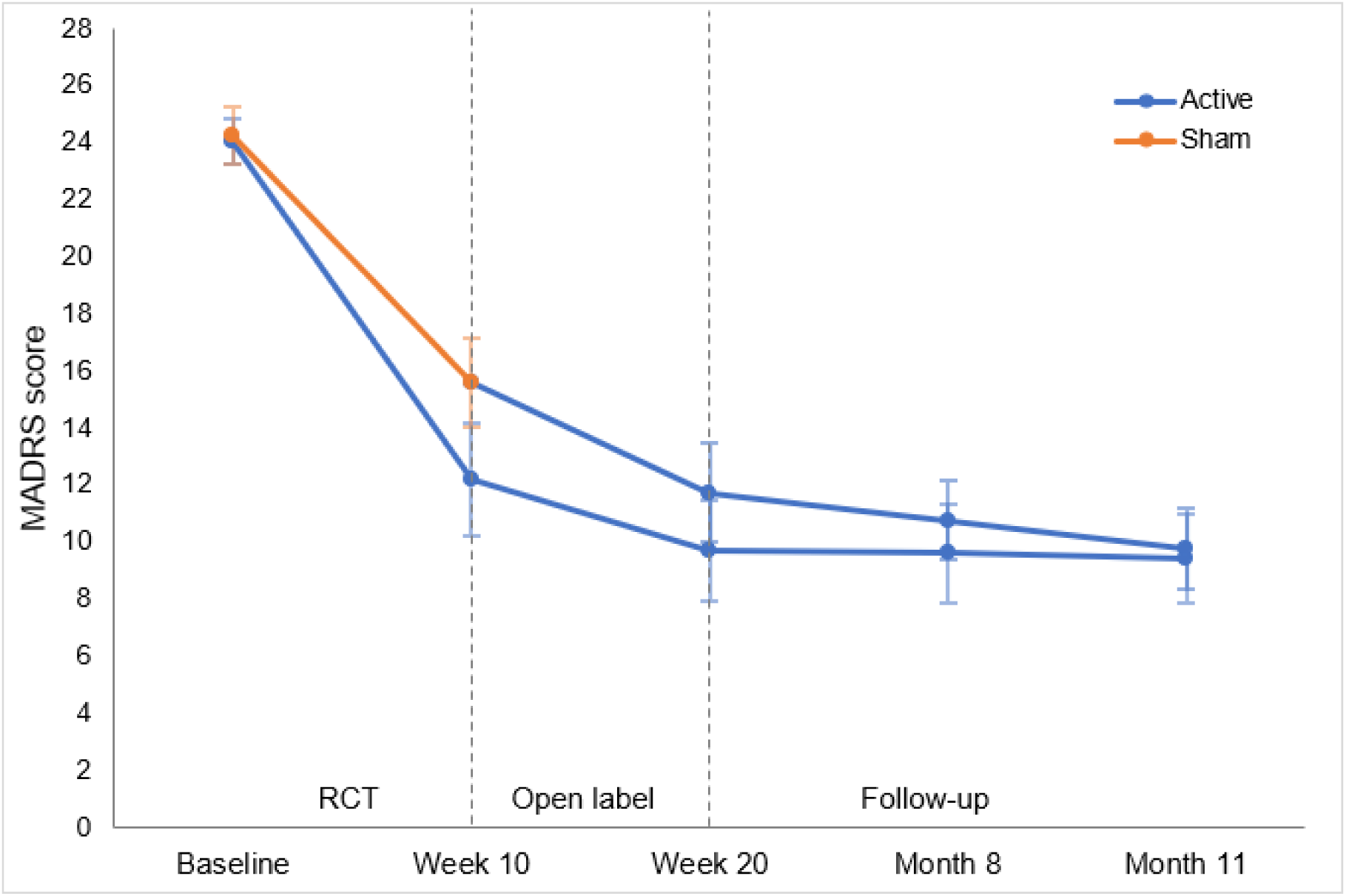
Montgomery-Åsberg Depression Rating Scale (MADRS) total scores for patients who were allocated to the active or sham treatment group at each assessment time point from the end of the blinded phase of the trial to the month 11 follow-up. Error bars represent standard deviation. Number of participants in the original active group (n=19) and in sham group (n=23).

**Figure 4.**
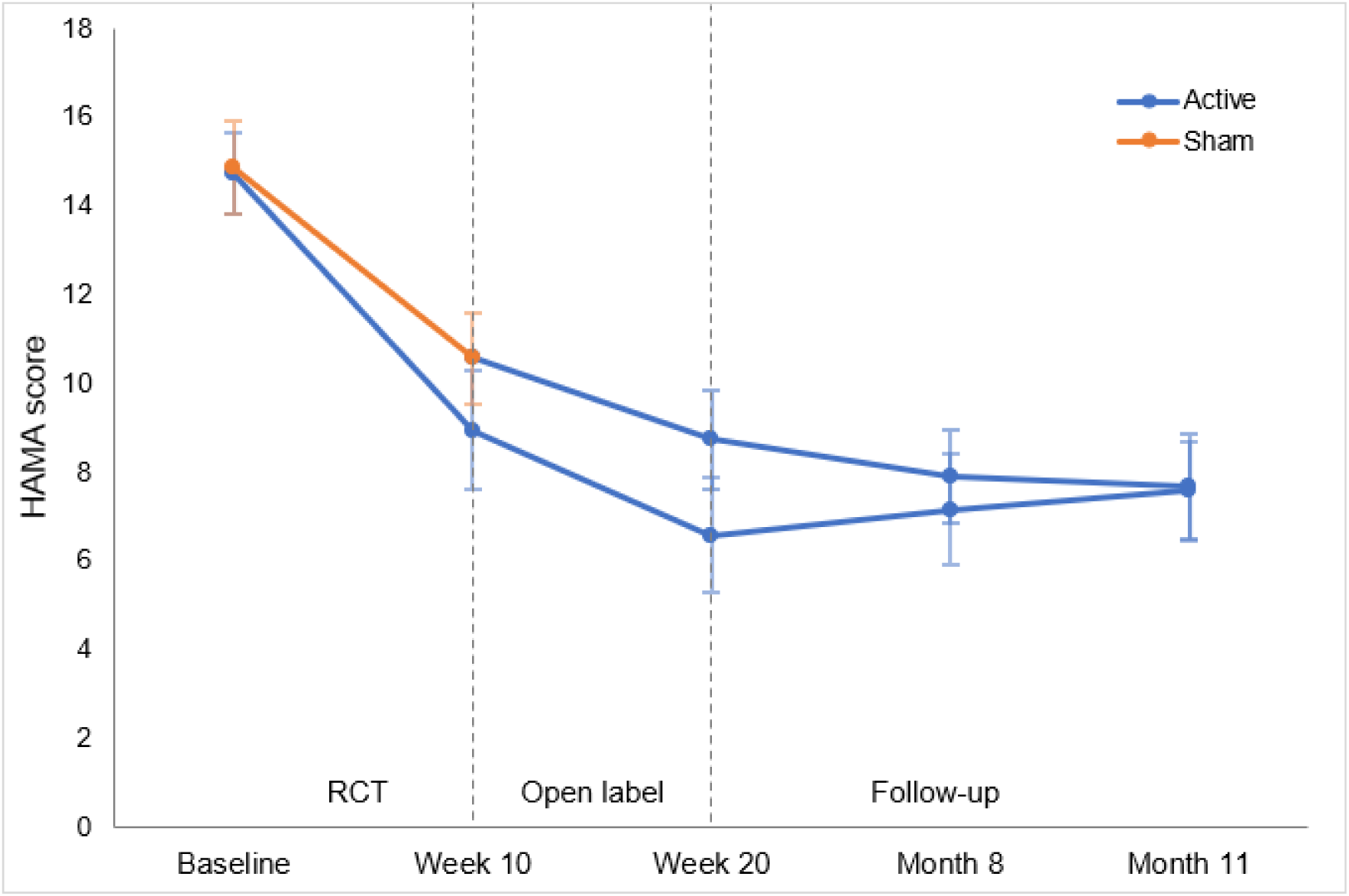
Mean Hamilton Anxiety Rating Scale (HAMA) total scores for patients who were allocated to the active or sham treatment group at each assessment time point from the end of the blinded phase of the trial to the month 11 follow-up. Error bars represent standard deviation. Number of participants in the original active group (n=19) and in sham group (n=23).

**Figure 5.**
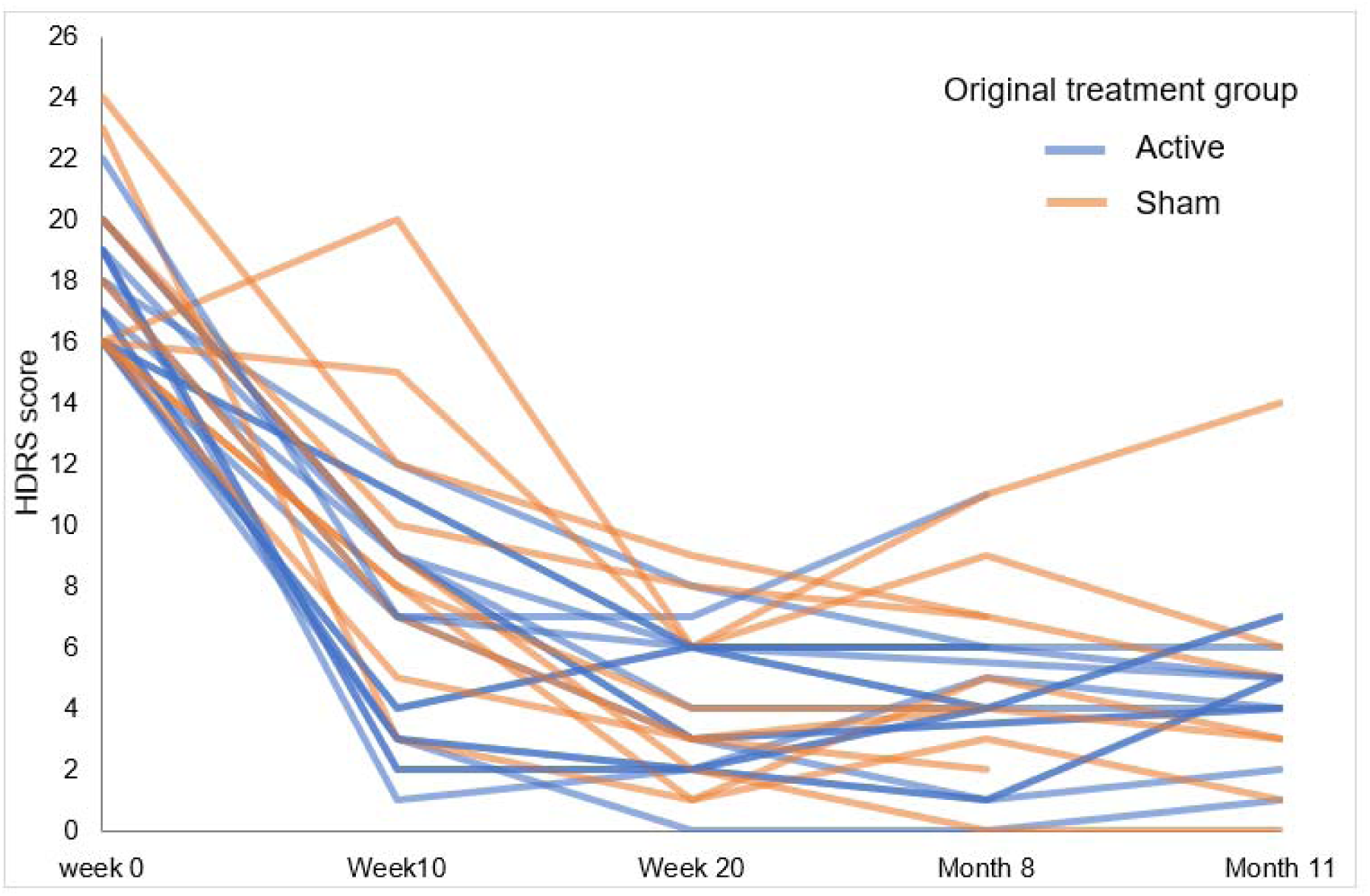
Hamilton Depression Rating Scale (HDRS) total scores for patients who showed a HDRS clinical response at the end of the acute phase of the trial at each follow-up time point (n=28).

Analyses to compare participants who had continued tDCS during the follow-up period with those who had not continued did not show any significant interaction effect between tDCS use during the follow-up period and time, at month 8 (*F*(1, 37) = 0.15, *p* = 0.71) nor at month 11 (*F*(1, 28) = 0.89, *p* = 0.35).

### 3.3. Outcomes for participants who showed a clinical response at week 20

From participants who showed clinical response at week 20 (n = 28) as measured by HDRS ratings, at month 8, participants completing follow-up (n = 26) showed the following clinical outcomes: clinical response (n = 22; 84%), remission (n = 21, 81%), and none had a depressive relapse. At month 11, of the participants completing follow-up (n = 21), (n = 19) participants showed clinical outcomes of both clinical response and remission (90%), one participant did not show a clinical response (5%) and one participant had a depressive relapse (5%) as measured by HDRS score of at least 14. tDCS device use patterns were: regular use over 6-month period (n = 10), use in first 3-month period only (n = 3), intermittent use over 6-month period (n = 5), and discontinued use (n = 3).

### 3.4. Safety and tolerability

Most common side effects were tingling, skin redness, itching and burning sensation, with less common reports of headache, scalp pain, acute mood change and sleepiness (Table 4). The severity was rated at mild: 85%, moderate: 13%, and severe: 2%, which were one report each for skin redness and acute mood change. There were no episodes of hypomania or mania as measured by YMRS scale and there were no suicide attempts as measured by C-SSRS.

**Table 4.**
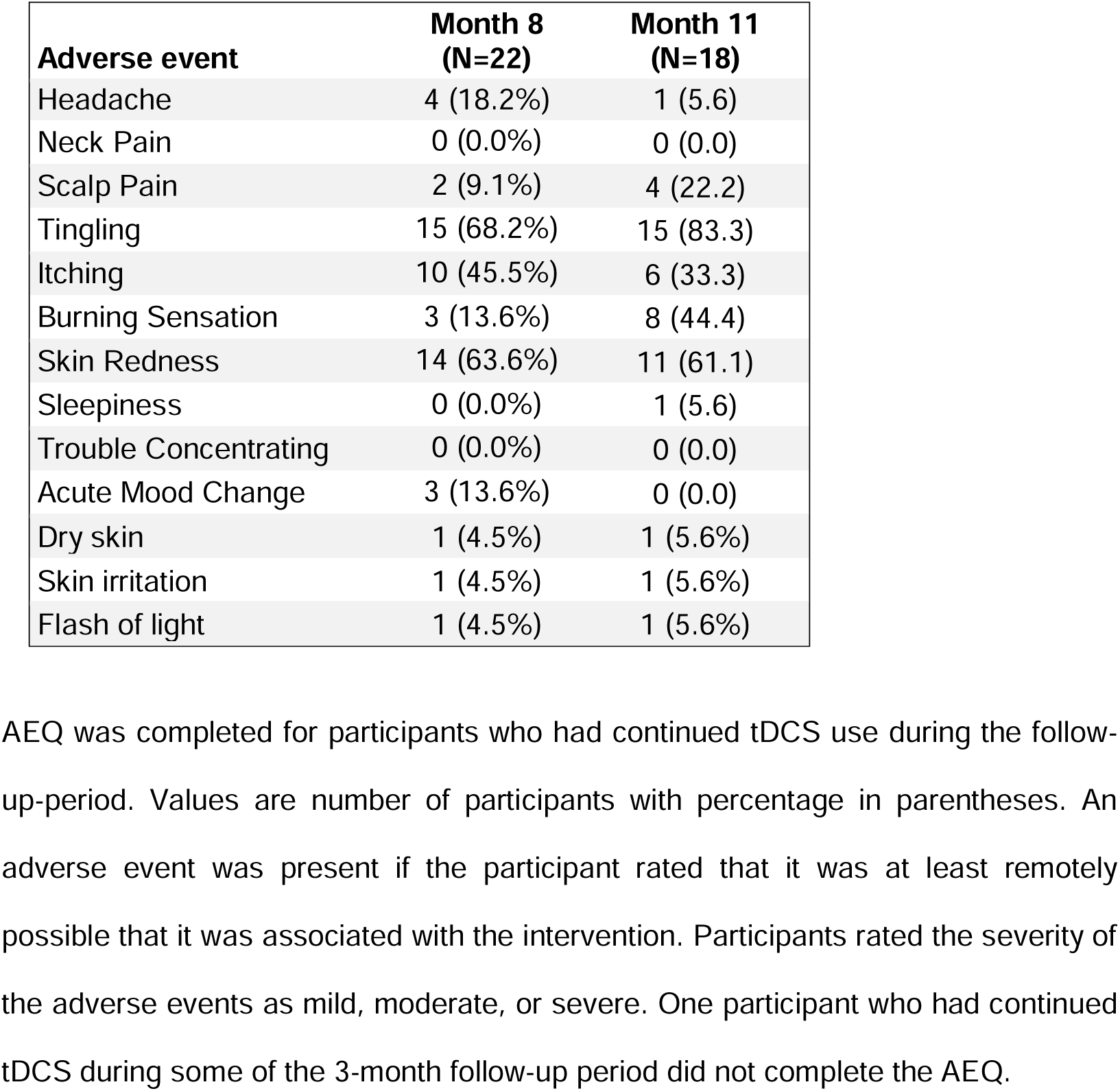
Adverse events at months 8 and 11 as measured by the tDCS Adverse Events Questionnaire. (Brunoni et al., 2011)

| **Adverse event** | **Month 8**<br>**(N=22)** | **Month 11**<br>**(N=18)** |
| --- | --- | --- |
| Headache | 4 (18.2%) | 1 (5.6) |
| Neck Pain | 0 (0.0%) | 0 (0.0) |
| Scalp Pain | 2 (9.1%) | 4 (22.2) |
| Tingling | 15 (68.2%) | 15 (83.3) |
| Itching | 10 (45.5%) | 6 (33.3) |
| Burning Sensation | 3 (13.6%) | 8 (44.4) |
| Skin Redness | 14 (63.6%) | 11 (61.1) |
| Sleepiness | 0 (0.0%) | 1 (5.6) |
| Trouble Concentrating | 0 (0.0%) | 0 (0.0) |
| Acute Mood Change | 3 (13.6%) | 0 (0.0) |
| Dry skin | 1 (4.5%) | 1 (5.6%) |
| Skin irritation | 1 (4.5%) | 1 (5.6%) |
| Flash of light | 1 (4.5%) | 1 (5.6%) |

AEQ was completed for participants who had continued tDCS use during the follow-up-period. Values are number of participants with percentage in parentheses. An adverse event was present if the participant rated that it was at least remotely possible that it was associated with the intervention. Participants rated the severity of the adverse events as mild, moderate, or severe. One participant who had continued tDCS during some of the 3-month follow-up period did not complete the AEQ.

At both month 8 and 11 follow-ups, acceptability was endorsed as being “very acceptable”, ethicality remained high at “very ethical”, effort required remained consistent at “the same amount of effort as usual”, impact of side effects was rated as “quite unaffected”, and participants “would strongly recommend” tDCS treatment to others. Ratings for perceived effectiveness were “quite helpful” at month 8 and “very helpful” at month 11 follow-up (Table 5).

**Table 5.** Acceptability questionnaire and responses at month 8 and month 11 follow-up.

| Question |  | Median (IQR) | Likert Ratings |  |  |  |  |  |  |
| --- | --- | --- | --- | --- | --- | --- | --- | --- | --- |
|  |  |  | 1 | 2 | 3 | 4 | 5 | 6 | 7 |
| How acceptable did you find the tDCS sessions? |  |  | Very unacceptable | Quite unacceptable | Unacceptable | Neither | Acceptable | Quite acceptable | Very acceptable |
|  | Month 8 | 7 (7, 7) | 0 (0%) | 0 (0%) | 1 (3%) | 0 (0%) | 0 (0%) | 8 (21%) | 29 (76%) |
|  | Month 11 | 7 (6, 7) | 0 (0%) | 0 (0%) | 0 (0%) | 0 (0%) | 0 (0%) | 9 (27%) | 24 (73%) |
| How helpful do you think the tDCS sessions were for improving your depressive symptoms? |  |  | Very unhelpful | Quite unhelpful | Bit unhelpful | Neither | Bit helpful | Quite helpful | Very helpful |
|  | Month 8 | 6 (6, 7) | 0 (0%) | 2 (5%) | 0 (0%) | 2 (5%) | 4 (11%) | 14 (37%) | 16 (42%) |
|  | Month 11 | 7 (6, 7) | 0 (0%) | 1 (3%) | 0 (0%) | 3 (9%) | 1 (3%) | 11 (33%) | 17 (52%) |
| How were you bothered by any negative side effects from the tDCS sessions? |  |  | Very much unaffected | Quite unaffected | Bit unaffected | Neither | Bit affected | Quite affected | Very affected |
|  | Month 8 | 2 (1, 5) | 17 (45%) | 6 (16%) | 1 (3%) | 3 (8%) | 10 (26%) | 1 (3%) | 0 (0%) |
|  | Month 11 | 2 (1, 4) | 14 (43%) | 8 (24%) | 1 (3%) | 2 (6%) | 8 (24%) | 0 (0%) | 0 (0%) |
| How ethical do you think the tDCS sessions are? |  |  | Very unethical | Quite unethical | Bit unethical | Neither | Bit ethical | Quite ethical | Very ethical |
|  | Month 8 | 7 (7, 7) | 0 (0%) | 0 (0%) | 0 (0%) | 0 (0%) | 1 (3%) | 4 (11%) | 33 (87%) |
|  | Month 11 | 7 (7, 7) | 0 (0%) | 0 (0%) | 0 (0%) | 1 (3%) | 0 (0%) | 6 (9%) | 29 (88%) |
| How much effort did you need to put in for the tDCS sessions? |  |  | Very much more than usual | Some more than usual | Little bit more than usual | Same as usual | Little bit less than usual | Some less than usual | Very much less than usual |
|  | Month 8 | 4 (3,5) | 0 (0%) | 4 (10%) | 13 (34%) | 10 (26%) | 3 (8%) | 3 (8%) | 5 (13%) |
|  | Month 11 | 4 (3,6) | 0 (0%) | 3 (9%) | 8 (24%) | 8 (24%) | 4 (12%) | 5 (15%) | 5 (15%) |
| Would you recommend the tDCS sessions to others? |  |  | Would very strongly not recommend | Would strongly not recommend | Would not recommend | Would not for or against | Would recommend | Would strongly recommend | Would very strongly recommend |
|  | Month 8 | 6 (6, 7) | 0 (0%) | 0 (0%) | 0 (0%) | 0 (0%) | 5 (13%) | 15 (40%) | 18 (47%) |
|  | Month 11 | 6 (6, 7) | 0 (0%) | 0 (0%) | 0 (0%) | 0 (0%) | 5 (15%) | 10 (30%) | 18 (55%) |

IQR, inter quartile range; tDCS, transcranial direct current stimulation. Month 8, n=38 (one participant did not complete the questionnaire); month 11, n=33.

## 4. Discussion

Long term follow-up assessments in participants from a phase 2 randomised controlled trial of home-based tDCS treatment of MDD demonstrated significant maintenance of treatment effects. All participants had engaged in the open-label phase of the trial and had received active tDCS (Woodham et al., 2024). Maintenance of treatment effects were observed at months 8 and 11 following randomisation, which was evident in high response and remission rates. In particular, participants who had a clinical response to at the end of the week 20 open label phase, 90% maintained remission at month 11.

Over half of participants continued to use the tDCS device in the follow-up period. No significant differences were found in depressive symptom scores between those who had continued and those who had not continued with tDCS use. Open-label studies of home-based tDCS treatment have observed continued high response and remission rates at 6 months (Alonzo et al., 2019; Woodham et al., 2022). Razza et al. (2021) meta-analysis treatment identified that interventional follow-up periods may lead to the continuation of improvements beyond the acute phase of treatment. In clinic-based studies, Aparicio et al. (2019) found relapse rates to be lower in an interventional study of participants who had responded to active tDCS, with a maintenance stimulation schedule of 2 times per week, compared to two interventional studies with less intensive maintenance stimulation schedules (Martin et al., 2013; Valiengo et al., 2013), indicating that more frequent schedules might be more beneficial.

Although our follow-up study was not interventional, we were able to consider the effects of continued tDCS treatment as many participants had chosen to continue with stimulation. When only considering the sub-sample participants who had responded to tDCS at the end of the 10-week open-label phase, response rates at months 8 and 11 were 84% and 90% respectively, which was higher than for the follow-up sample as a whole which was 64% and 76% respectively. The majority of participants who had maintained clinical response from the end of the 20-week open label phase to the month 11 follow-up had continued with tDCS use. In addition to exploring the frequency of follow-up maintenance stimulation, future studies with interventional follow-up periods might aim to compare continued maintenance stimulation and no stimulation to better understand if relapse rates are comparable.

The continued use of tDCS by participants in long term follow-up further demonstrates the acceptability and feasibility of home-based self-administered tDCS over a longer time period. Although tDCS use was not being monitored by the study team, acceptability reports remained high, no serious adverse events occurred, and no participants developed mania or hypomania.

Relapse following rTMS treatment is more likely in the absence of maintenance treatment. Interventional maintenance periods are effective for continued clinical response beyond the acute treatment phase and in preventing relapse (Brian Chen et al., 2023; Matsuda et al., 2023). However, continued treatment requires regular visits to clinics. rTMS and tDCS are comparable in their ability to reduce electrophysiological complexity in MDD (Čukić, 2019). Future research could explore the effectiveness of home-based tDCS as a more accessible maintenance treatment for rTMS and other treatments. A clinical question of significance is whether continued maintenance tDCS sessions beyond the acute treatment phase is necessary or beneficial for preventing future relapse. Due to the portability and low cost, continued maintenance treatment could be feasible, as is typically the case with antidepressant medication which is maintained in order to prevent a relapse.

Limitations of the present study include being a sub-sample of the original RCT sample, which limited power to detect group differences. The observational design of the study did not control for frequency of tDCS use, nor adherence to a specific schedule, therefore analyses comparing those who had continued stimulation with those who had not, have limited clinical implications. Not all participants continued in the long term follow-up. The discontinuation rate was 21% at the month 11 follow-up, which is comparable to long term tDCS follow-up studies (Aparicio et al., 2019; Martin et al., 2013; Valiengo et al., 2013). The sample included participants who had shown a treatment response (67%) and those who had not (33%) at the end of the week 10 RCT, therefore, samples comparing groups likely include participants for whom tDCS is not effective. Future interventional studies with control groups are necessary to better understand the role of maintenance tDCS in the year following acute treatment and to help determine the most effective schedule of maintenance treatment. Our sample was predominantly participants of white ethnicity, and exclusion criteria included treatment resistant depression and history of hospital admissions which may limit the generalisability of the findings.

## 5. Conclusion

In conclusion, long term response and remission rates were maintained at months 8 and 11 following a 10-week RCT and 10-week open label treatment. In particular, participants who had shown a clinical response at the end of the 20-week open label phase, the majority maintained a clinical response and remission. No significant differences were found in depressive symptoms between those who had continued tDCS with those who had not in the long term. Over half of participants chose to continue with tDCS in the long term, indicating that long-term home-based tDCS is acceptable and feasible.

## Ethics statement

All procedures involving human subjects/patients were approved by research ethics boards of the University of East London and by South Central-Hampshire B Research Ethics Committee, UK (ref. 22/SC/0023). Study procedures were explained to participants prior to signing informed consent. All procedures contributing to this work comply with the ethical standards of the relevant national and institutional committees on human experimentation and with the Helsinki Declaration of 1975, as revised in 2008.

## Data Statement

Anonymised data will be made available on request.

## Author contributions – CRediT

**Rachel Woodham:** Data Curation, Investigation, Formal Analysis, Writing-Original draft preparation, Writing-Reviewing and Editing; **Sudhakar Selvaraj:** Project administration, Writing-Reviewing and Editing. **Nahed Lajmi:** Investigation, Writing-Reviewing and Editing. **Harriet Hobday:** Investigation, Writing-Reviewing and Editing. **Gabrielle Sheehan:** Investigation, Writing-Reviewing and Editing. **Ali-Reza Ghazi-Noori:** Investigation, Writing-Reviewing and Editing. **Peter Lagerberg:** Investigation, Writing-Reviewing and Editing. **Rodrigo Machado-Vieira:** Project administration, Writing-Reviewing and Editing. **Jair Soares:** Methodology, Writing-Reviewing and Editing. **Allan H. Young:** Methodology, Writing-Reviewing and Editing. **Cynthia H.Y. Fu:** Conceptualization, Methodology, Funding Acquisition, Project Administration, Supervision, Writing-Original draft preparation, Writing-Reviewing and Editing.

## Acknowledgements

We would like to acknowledge the participants for their contribution to the study, and research assistants Maheen Rizvi, Paulette Orhii and Sarah Kwon for their support with data acquisition in the RCT. Professor Young’s independent research is funded by the National Institute for Health and Care Research (NIHR) Maudsley Biomedical Research Centre at South London and Maudsley NHS Foundation Trust and King’s College London. The views expressed are those of the author(s) and not necessarily those of the NIHR or the Department of Health and Social Care.

